# Outcomes of COVID-19: disparities by ethnicity

**DOI:** 10.1101/2020.10.11.20210740

**Authors:** Hamad Ali, Abdullah Alshukry, Sulaiman K Marafie, Monera AlRukhayes, Yaseen Ali, Mohammad Bu Abbas, Abdullah Al-Taweel, Yousef Bukhamseen, Mohammad H Dashti, Abdullah Al-Shammari, Mohammad Abu-Farha, Jehad AbuBaker, Fahd Al-Mulla

## Abstract

**Objectives:** To investigate the role of ethnicity in COVID-19 outcome disparities in a cohort in Kuwait.

**Methods:** This is a retrospective analysis of 405 individuals infected with SARS-CoV-2 in Kuwait. Outcomes such as symptoms severity and mortality were considered. Multivariate logistic regression models were used to report the odds ratios (OR) for ICU admission and dying from COVID-19.

**Results:** The cohort included 290 Arabs and 115 South Asians. South Asians recorded significantly higher COVID-19 death rates compared to Arabs (33% vs. 7.6%, P value<0.001). When compared to Arabs, South Asians also had higher odds of being admitted to the ICU (OR = 6.28, 95% CI: 3.34 – 11.80, p < 0.001). South Asian patients showed 7.62 (95% CI: 3.62 – 16.02, p < 0.001) times the odds of dying from COVID-19.

**Conclusion:** COVID-19 patients with South Asians ethnicity are more likely to have worse prognosis and outcome when compared to patients with Arab ethnicity. This suggest a possible role for ethnicity in COVID-19 outcome disparities and this role is likely to be multifactorial.

## Introduction

Clinical manifestations of COVID-19 patients vary broadly, ranging from asymptomatic infection to acute respiratory failure and death (Alshukry et al., 2020). It is well established now that age, gender and certain pre-existing health conditions are considered as risk factors for a severe form of the disease (Ali et al., 2020). However certain indications have suggested a possible implication for ethnicity and race as contributors to the wide disparity in COVID-19 outcomes. Recent findings in the United States and United Kingdom showed differences in COVID-19 related hospitalization and death rates by ethnicity (Garg et al., 2020; Patel et al., 2020). Ethnicity is influenced by genetic backgrounds, environmental factors, as well as cultural and behavioral norms. Others have linked ethnicity to socioeconomic status, living standards and occupations (Patel et al., 2020). These factors linked to ethnicity are likely to function interdependence and influence disease outcome. As COVID-19 continue to spread globally, understanding how ethnicity affects SARS-CoV2 susceptibility and disease outcome would surely improve disease management and outcome.

Kuwait is a multiethnic country with a population of 4.3 million. Arabs make 59.2% of the population while South Asians make 37.8% (Central Statistical Bureau of Kuwait (CSBoK), 2019). In here, we present and compare COVID-19 clinical outcomes in the Arabs and South Asians ethnic groups.

## Material and Methods

### Cohort details

Four hundred and five consecutive COVID-19 patients were recruited in this study. Patients were admitted to Jaber Al-Ahmad Hospital in Kuwait between February 24 and May 24, 2020. Hospitalization rate was 100%, subject to a positive PCR test regardless of disease severity. Cohort clinical characteristics were previously described by our group (Ali et al., 2020; Alshukry et al., 2020). The study was reviewed and granted an approval by the standing committee for coordination of health and medical research in the Ministry of Health in Kuwait (IRB 2020/1404). Patients were divided into two main groups; Arabs and South Asian. Disease severity was determined following criteria described previously (Alshukry et al., 2020).

### Statistical analysis

Student T-test was used to examine continuous variables, results were reported as mean ± standard deviation. Nominal variables were determined by using χ2 test and categorical variables compared using Fisher-Exact test. We investigated the relationships between ethnicity as a categorical exposure and both (1) the death outcome from COVID-19 and (2) ICU admission. We used binary response variables and adjusted for age, gender, smoking status and other co-morbidities. A logistic regression model was used to report the odds ratios (OR) for dying from COVID-19 and for ICU admission for patients of South Asian ethnicity, with a baseline comparison to Arab ethnicity.

## Results

Arabs made the majority of the cohort (71.6%). Males were predominant in both ethnic groups which were also age matched. Mortality rate in Arabs groups was 7.5% while it reached 33% in the South Asians (p<0.001). No significant differences were reported in comorbidities prevalence between both groups (Table.1). South Asians also had higher odds of being admitted to the ICU (OR = 6.28, 95% CI: 3.34 – 11.80, p < 0.001). (Figure.1). They also showed 7.62 (95% CI: 3.62 – 16.02, p < 0.001) times the odds of dying from COVID-19 when compared to Arabs (Figure.1).

**Table 1.** Cohort characteristics

|  | Arabs |  | South Asians |  |  |
| --- | --- | --- | --- | --- | --- |
|  | count | % | count | % | P value |
|  | 290 | 71.6 | 115 | 28.4 |  |
| **Gender** |  |  |  |  |  |
| Male | 164 | 56.6 | 91 | 79.1 | <0.001 |
| Female | 126 | 43.4 | 24 | 20.9 | <0.001 |
| Age | (Mean years ± SD) | | (Mean years ± SD) | | 0.069 |
| | 44.5 ± 18.2 | | 47.6 ± 13.4 | | |
| **Disease spectrum** |  |  |  |  |  |
| Asymptomatic | 136 | 46.9 | 25 | 21.7 | <0.001 |
| Mild/Moderate | 119 | 41.0 | 43 | 37.4 | 0.499 |
| ICU | 35 | 12.1 | 49 | 42.6 | < 0.001 |
| Deaths | 22 | 7.6 | 38 | 33 | < 0.001 |
| **Comorbidities** |  |  |  |  |  |
| Diabetes | 67 | 23.3 | 30 | 26.1 | 0.606 |
| Hypertension | 93 | 32.2 | 28 | 24.3 | 0.148 |
| Asthma | 31 | 10.8 | 9 | 7.9 | 0.462 |
| Cardiovascular disease | 31 | 10.7 | 8 | 7.0 | 0.350 |
| Chronic renal disease | 12 | 4.2 | 2 | 1.7 | 0.367 |
| Malignancy | 9 | 3.1 | 3 | 2.6 | 0.784 |

**Figure 1.**
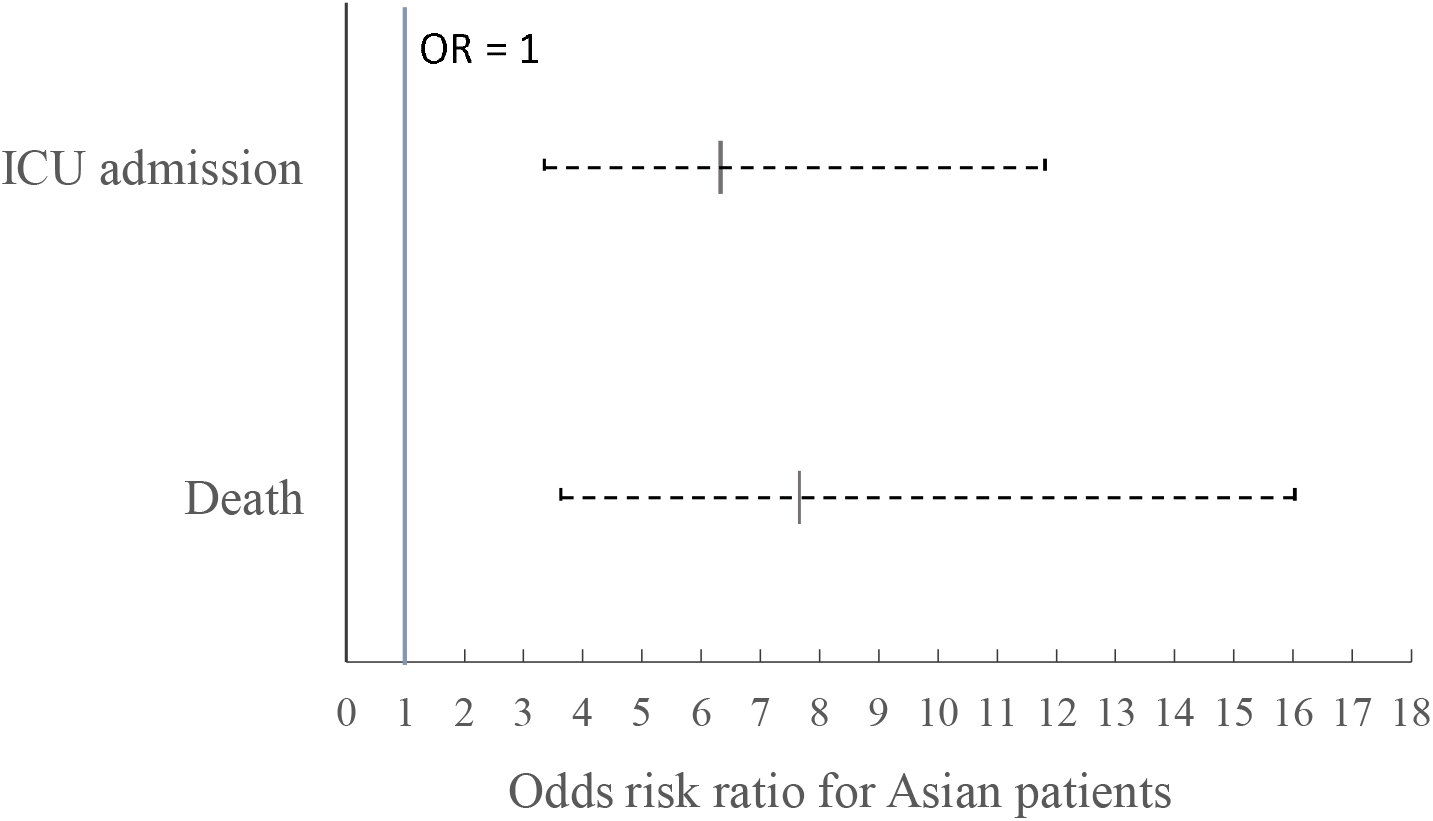
Odds ratios of dying and being admitted to ICU from COVID-19 if a patient is of South Asian ethnicity. Baseline is Arab ethnicity. ORs are adjusted for age, gender, smoking status and other comorbidities.

## Discussion

Results from our cohort, which provided good representation of the population, indicated that South Asians were more likely to develop a severe form of the disease which also corresponded to a higher death rate compared to Arabs.

Our results suggested ethnicity as a possible risk factor for COVID-19 poor prognosis and outcome. The vast majority of South Asians in Kuwait are unskilled labor living in highly populated accommodations. Their living conditions and perhaps occupation can increase their exposure to SARS-CoV2. While this could have a major impact on infection dynamics, it may not explain the results entirely. Local laws grant each resident in Kuwait free access to governmental health care, hence rolling out healthcare access inequalities of such outcome disparity. Comorbidities are known as risk factors for severe and poor outcomes however our results indicated no significant differences in prevalence between the two ethnic groups and therefore do not provide a conclusive answer for the disparity in outcome. We certainly cannot overlook the role of genetic factors which might make people from certain ethnicity more prone to severe outcome (Al-Mulla et al., 2020).

The role of ethnicity in COVID-19 outcome disparities is likely to be multifactorial in nature. Understanding why certain ethnic groups are prone to severe COVID-19 outcome is important to improve outcomes and develop effective mitigation strategies.

## Data Availability

The dataset generated during and/or analyzed during the current study is available in the Figshare repository.

https://doi.org/10.6084/m9.figshare.12567881.v1

## Authors contribution

HA involved in the drafted the article and had the final approval of the version to be submitted. AA1, YA, MB, AA2, YB and MD are involved in acquisition of the data. SM, MA and AA3 are involved in data analysis. MA and JA involved in data interpretation. FA revised the article critically for important intellectual content.

## Ethical approval

The study was reviewed and granted an approval by the standing committee for coordination of health and medical research in the Ministry of Health in Kuwait (IRB 2020/1404).

